# Lockdowns and Selection Pressure: A Modelling Study

**DOI:** 10.1101/2023.06.13.23291336

**Authors:** James Thompson, Stephen Wattam

**Affiliations:** Department of Infectious Disease Epidemiology, Imperial College London; WAP Academic Consultancy

## Abstract

Since COVID-19 was first identified in China in 2019, the SARS-CoV-2 virus has mutated and given rise to a large number of variants. A range of non-pharmaceutical interventions have been deployed against the virus in an effort to save lives and reduce pressure on health-care systems. These interventions may have favoured some variants over others. Despite this possibility, there has thus far been very little work investigating the impact of such interventions on the evolution of the virus.

Using mathematical and computational models, we investigate the impact of a lockdown specifically, for the case in which two SARS-CoV-2 variants are circulating simultaneously in a population. We find that under certain conditions, lockdowns disrupt the competition between variants in such a way that highly transmissible variants with long infectious periods are selected for, ultimately leading to *more* cases overall than would have occurred without the lockdown, due to larger second waves of cases.

These are the results of a modelling study and we do not claim to have found evidence of such unfavourable selection effects occurring in reality. On the other hand, our results are consistent with evolutionary theory and suggest that the selective effects of non-pharmaceutical interventions deserve greater scrutiny.

## Introduction

The first COVID-19 lockdown was implemented in Wuhan, on 23rd January 2020, resulting in travel restrictions, non-essential business closures and stay-at-home orders for all 11 million inhabitants of the city. Similar restrictions were soon after imposed in many other parts of China and across the world. These lockdowns were of unprecedented length, lasting in some cases many months. While a lockdown was implemented in Mexico in 2009, in response to the swine flu outbreak [4], and another in Sierra Leone in 2015, in response to the Ebola outbreak [5, 17], these lockdowns lasted only 5 and 3 days, respectively. Even during the 1918-1919 influenza pandemic, restrictions on mobility in cities across the United States were less stringent than those imposed by the COVID-19 lockdowns [13]. Moreover, literature on lockdowns dating from before the COVID-19 pandemic is extremely limited. For example, searching PubMed and the preprint servers medRxiv and bioRxiv for items containing the keyword ‘lockdown’ yields 24375 results at the time of writing, but only 89 of these results date from 2019 or earlier, with only 1 result being related to the control of infectious diseases. Pandemic preparedness studies typically considered social distancing interventions far less stringent than the COVID-19 lockdowns [1, 10, 11, 14], due in part to ethical considerations [3]. The COVID-19 lockdowns were, therefore, implemented without precedent and before a deep scientific understanding of their impact and long-term consequences had been established.

Social interaction is reduced for the duration of a lockdown, thereby reducing transmission and temporarily easing pressure on healthcare systems. On the other hand, lockdowns incur substantial social and economic costs. In this article we ignore such costs, focussing instead on the claimed public health benefit of lockdowns against an evolving virus such as SARS-CoV-2.

Very little attention has been given to the evolutionary impact of non-pharmaceutical interventions in general. Recent works addressing this topic in the context of the COVID-19 pandemic include the articles of Gurevich et al. [7], Ashby and Thompson [2] and Nielsen et al. [15]. Gurevich et al. [7] examined the effect of testing and isolation on the evolution of SARS-CoV-2, finding that testing and isolation may act to reduce virulence. They found that a higher testing rate can select for a test-evasive viral strain, even if that strain is less infectious than the competing detectable strain. Ashby and Thompson [2] argued that while stronger and timelier non-pharmacheutical interventions generally reduce the likelihood of variant emergence, it is possible for more transmissible variants to have a greater probability of emerging at intermediate levels of these interventions. Nielsen et al. [15] considered the impact of lockdowns specifically, and found that lockdowns exert an evolutionary pressure which favours variants with lower levels of overdispersion.

We will investigate the impact of lockdowns using both an SIR model and an agent-based model. The equation-based SIR model is simple and offers generality, while the agent-based model is more detailed and realistic. The latter has been validated using COVID-19 clinical monitoring data collected in Luxembourg and is the model used previously by the authors in Thompson and Wattam [20].

In each model, we suppose that two viral strains are circulating simultaneously in a population. We assume that individuals cannot be simultaneously infected with both strains and that recovery from one strain implies immunity to both strains. The properties of the reference strain are determined using real-world data, while the properties of the alternative strain are determined by modifying those of the reference strain. For a range of such modifications, we simulate a lockdown and compare the final outcome to that of the baseline scenario in which no interventions are active. We verify using both the equation-based and agent-based modelling approaches and obtain consistent results.

Our modelling suggests that, in certain circumstances, a lockdown can disrupt the competition between strains in such a way that ultimately results in *more* cases than would have occurred without the lockdown. This effect can be understood in terms of *r/K* selection theory. With the strains engaged in an exploitative competition for hosts, the lockdown shifts the selection regime from *r*-selection, which favours high growth rates, to *K*-selection, which favours high carrying capacities. With limited access to hosts during a lockdown, strains with longer infectious periods are favoured, even at the expense of lower transmission probabilities. This may ultimately lead to more infections overall through larger second waves, since the strains selected for have a higher *R*_0_. These are only the results of a modelling study, so further investigation is required to determine if the lockdowns against COVID-19 did in fact cause or accelerate the selection of more transmissible variants.

## Materials and Methods

### SIR Model

The SIR model is an epidemic model based on homogeneous mixing. It is given by the system of ordinary differential equations

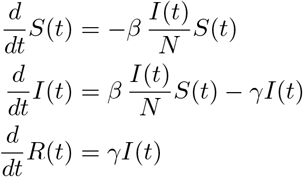

for which we set (*S*(0), *I*(0), *R*(0)) = (625640, 320, 0). Here *S*(*t*), *I*(*t*) and *R*(*t*) denote the number of individuals susceptible, infected and recovered, respectively, at time *t*, while *N* = *S*(*t*) + *I*(*t*) + *R*(*t*) denotes the total population size. We set *N* = 625960 since this is approximately the size of the resident population of Luxembourg, on which the agent-based model has been validated, and to which we will later compare the SIR model. We have set *I*(0) = 320 for similar reasons. The parameter *β* denotes the average contact rate multiplied by the transmission probability, while the parameter *γ* denotes the recovery rate. In the context of COVID-19, social distancing restrictions (which reduce the contact rate) and the wearing of face masks (which reduce the transmission probability) can be represented as temporary reductions in *β*.

Now let us introduce a second strain, referring to the two strains as **Strain 1** and **Strain 2**, and consider the following two-strain SIR model:

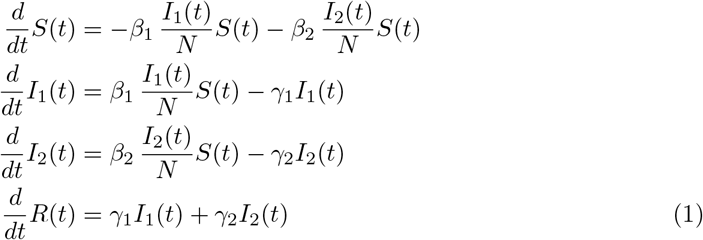

with initial conditions (*S*(0), *I*_1_(0), *I*_2_(0), *R*(0)) = (625320, 320, 320, 0). Here *I*_1_(*t*) and *I*_2_(*t*) denote the number of individuals infected with **Strain 1** and **Strain 2**, respectively, at time *t* The parameters *β*_1_ and *β*_2_ denote the transmission rates for the two strains, while *γ*_1_ and *γ*_2_ denote the recovery rates.

### Agent-Based Model

If the equation-based approach of the SIR model is *top-down*, then the agent-based approach is *bottom-up*. Together these two approaches provide a more complete analysis than would have been obtained using only one or the other. In an agent-based model, the simultaneous actions and interactions of multiple individuals are simulated in an attempt to re-create and predict the appearance of complex phenomena resulting from their collective behaviour. Such models have been used extensively to study the spread of infectious diseases such as COVID-19. The basic features of our model have already been described, in some detail, in Thompson and Wattam [20]. It is a highly heterogeneous stochastic model based on collocation, in which agents move between locations inside a procedurally generated random environment.

#### Code

The model is written mainly in Python, with the code being open-source and publicly available on GitHub. The original version can be found here:

https://github.com/abm-covid-lux/abmlux and the version of it modified to accommodate multiple strain can be found here:

https://github.com/abm-covid-lux/multi_strain_abmlux

A faster version of the latter, partially rewritten in C**++** and containing tools to generate each of the plots found in this article, can be found here:

https://github.com/abm-covid-lux/multi_strain_abmlux_fast

The file `config.yaml`, found in the `States` folder of the `multi_strain_abmlux_fast` repository, indicates precisely which values are taken by each of the parameters appearing in the model.

#### Model Description

We start with the following basic assumptions on human behaviour:

1. Individuals perform sequences of activities.
2. Individuals perform these activities at particular locations.

By an *activity* we mean anything that an individual *does*, for example cooking, driving to work, or shopping. By a *location* we simply mean somewhere such as a house, restaurant, shop or school classroom. Denoting by *A* the set of all activities, we assign to each individual *i* a sequence

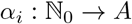

with *α*_*i*_(*t*) denoting the activity being performed by individual *i* at time *t*. Here N_0_ is the set of natural numbers 0, 1, 2, …. Denoting by *L* the set of all locations, we then assign to each individual *i* a map

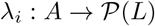

from *A* to the power set of *L*. By the power set of *L*, we mean the set of all subsets of *L*. For each activity, the map *λ*_*i*_ determines a set of locations at which individual *i* can perform that activity. It follows that *λ*_*i*_(*α*_*i*_(*t*)) is the set of locations at which individual *i* could be located at time *t*. Our model assumes that individuals choose from within this set uniformly at random whenever a change of activity occurs, with the maps *λ*_*i*_ being independent of *t* to ease the computational burden of the model.

We suppose that several strains of a virus are circulating in the population and that the health of an individual can be described by one of a finite number of health states. The health state of a individual is initially **Susceptible** and after infection ultimately becomes either **Recovered** or **Dead**, passing through a number of intermediate states along the way and spending a certain amount of time in each, according to a uniquely determined disease progression defined for that individual. Let us denote by *H* the set of all health states and by *S* the set of all strains. We then have a function

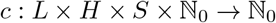

where *c*(*l, h, s, t*) is by definition the number of individuals in location *l* infected with strain *s* in health state *h* at time *t*.

To each location *l* we also assign a transmission probability multiplier *μ*(*l*) ϵ [0, 1], representing the fact that some locations are less conducive to disease transmission than others. For example, we suppose *μ*(*l*) = 0 for all outdoor locations. To each health state *h* and a strain *s* we assign a transmission probability *p*(*h, s*) ϵ [0, 1], such that if precisely two individuals are in location *l* at time *t*, with one individual susceptible and the other infected with strain *s* and in health state *h*, then *μ*(*l*)*p*(*h, s*) is the probability that the susceptible individual contracts the virus at time *t*. More generally, for any location *l*, the probability that a susceptible individual contracts the virus while in location *l* at time *t* is given by the expression

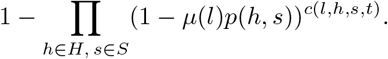

If such an individual contracts the virus, then to determine which strain that individual is infected with, we assume that the expression

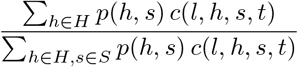

gives the probability that they are infected with strain *s*, for each *s ϵ S*.

#### Input Data

The agent-based model has been configured to represent the small western European country of Luxembourg, together with populations of cross-border workers in the neighbouring countries of Belgium, France and Germany. As described in [20], the sequences of activities *α* and location maps *λ* are configured using data collected by STATEC, the government statistics service of Luxembourg [18], and MMTP, the Ministry of Mobility and Public Transport of the government of Luxembourg, respectively. In particular, behavioural data comes from the 2014 Luxembourg Time Use Survey [19] while mobility data comes from the 2017 Luxmobil Survey [12]. Location data come from STATEC and OpenStreetMap [16]. Population grid data comes from the 2011 GEOSTAT study, organized by Eurostat [6]. Census data, collected by STATEC, was used to configure population age structure and household composition. COVID-19 clinical monitoring data comes from IGSS, the General Inspectorate of Social Security of Luxembourg [8]. We validated the model using data on COVID-19 hospitalizations and deaths in Luxembourg for the period February 2020 to July 2020. The reference strain appearing in our agent-based model therefore reflects the average of all SARS-CoV-2 variants circulating in Luxembourg during that specific period of time. Simulations begin with 320 randomly selected initial cases for each strain.

## Results

We first present the results of the SIR model, after which we verify them using the agent-based model.

### SIR Model

To assess the impact of a lockdown, we consider the following two scenarios:

- **Baseline:** No interventions are active.
- **Lockdown:** Contact rates are reduced by 90% for 28 days, starting on day 21.

We fix **Strain 1**, with a mean infectious period of 9 days and *R*_0_ = 2.45. In the two-strain SIR model, **Strain 1** is therefore specified by the parameters *β*_1_ = 0.2723 and 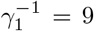 with the units of these parameters being days^*−*1^ and days, respectively. For **Strain 2**, we suppose *β*_2_ = *aβ*_1_ and *γ*_2_ = *b*^*−*1^*γ*_1_ for constants *a >* 0 and *b >* 0. Then *a* represents the *transmission probability ratio* and *b* the *infectious period ratio*. We allow *a* and *b* to vary over a range of values, given by a discretization of the square

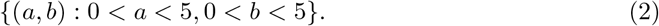

We set equal numbers of initial cases for each strain, and for each pair (*a, b*) we solve the two-strain SIR system (1) numerically using a forwards Euler scheme, over a period of 400 days, and calculate the final difference in *R* between the two scenarios. That is, we subtract the total number of recovered individuals after 400 days of the **Lockdown** scenario from the total number of recovered individuals after 400 days of the **Baseline** scenario. We plot these differences in Figure 1. If the difference is positive, then the lockdown has reduced total cumulative cases and we colour the square purple. If the difference is negative, then the lockdown has increased total cumulative cases, and we colour the square orange. The white squares indicate regions of the parameter space in which the difference is close to zero.

**Figure 1:**
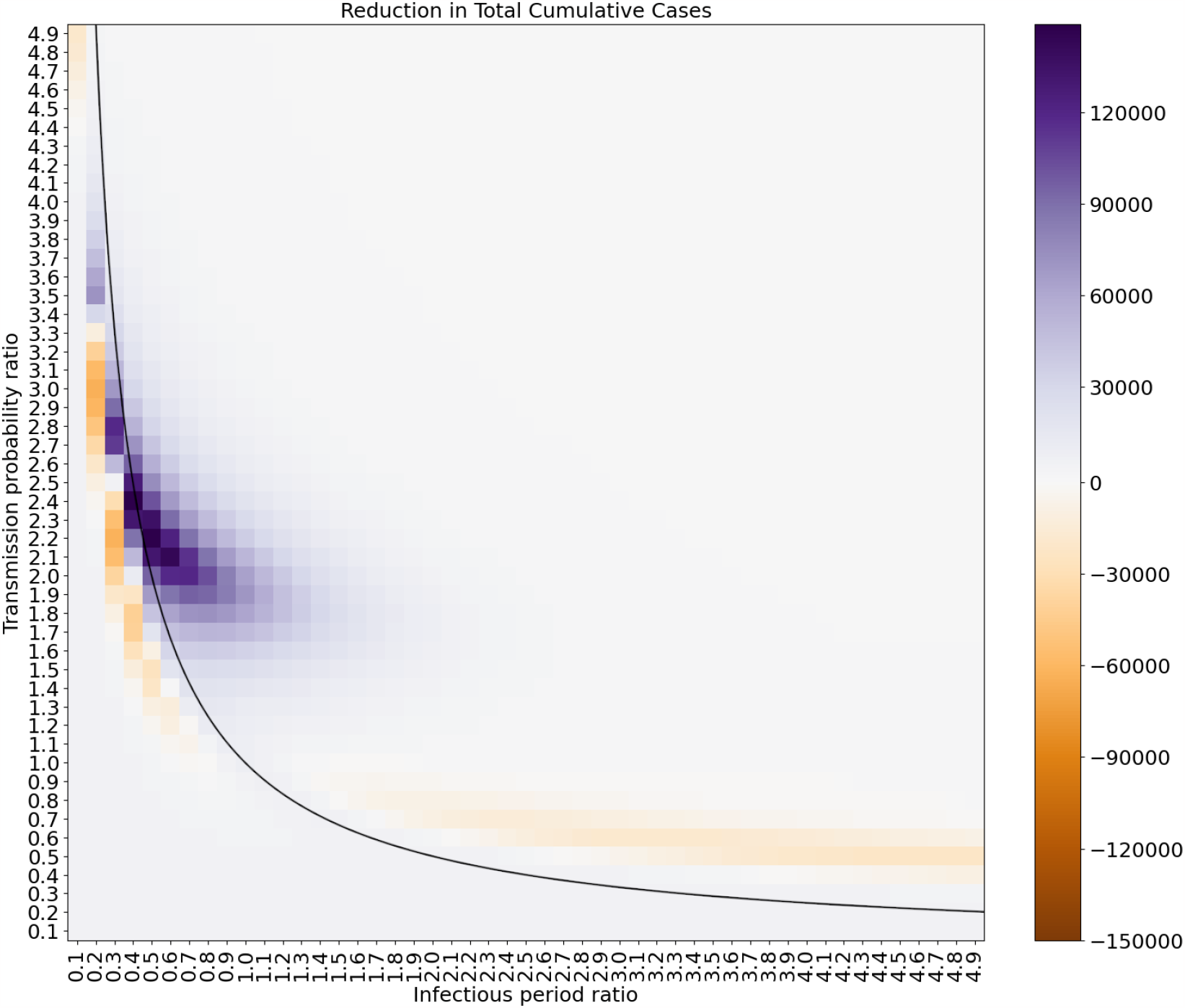
The impact of a lockdown in the two-strain SIR model. The thin orange regions are subsets of the parameter space in which the lockdown results in more cases than would have occurred without the lockdown. On the plotted curve *a* = 1*/b* the *R*_0_’s of the two strains are identical. At the point (1.0, 1.0), the strains themselves are identical.

For the region above the curve *a* = 1*/b* in Figure 1, **Strain 2** has the higher *R*_0_, due to its longer infectious period outweighing its smaller transmission probability. For the orange region below the curve *a* = 1*/b*, the same is true of **Strain 1**. Let us consider for example the pair (*a, b*) = (3.0, 0.2). In this case, **Strain 2** has a transmission probability 3 times that of **Strain 1**, while **Strain 1** has an infectious period 5 times that of **Strain 2**, meaning that **Strain 1** has a larger basic reproduction number *R*_0_ than **Strain 2**. In Figure 2, we plot daily cases and cumulative cases for these two strains, with **Strain 1** coloured red and **Strain 2** coloured blue.

**Figure 2:**
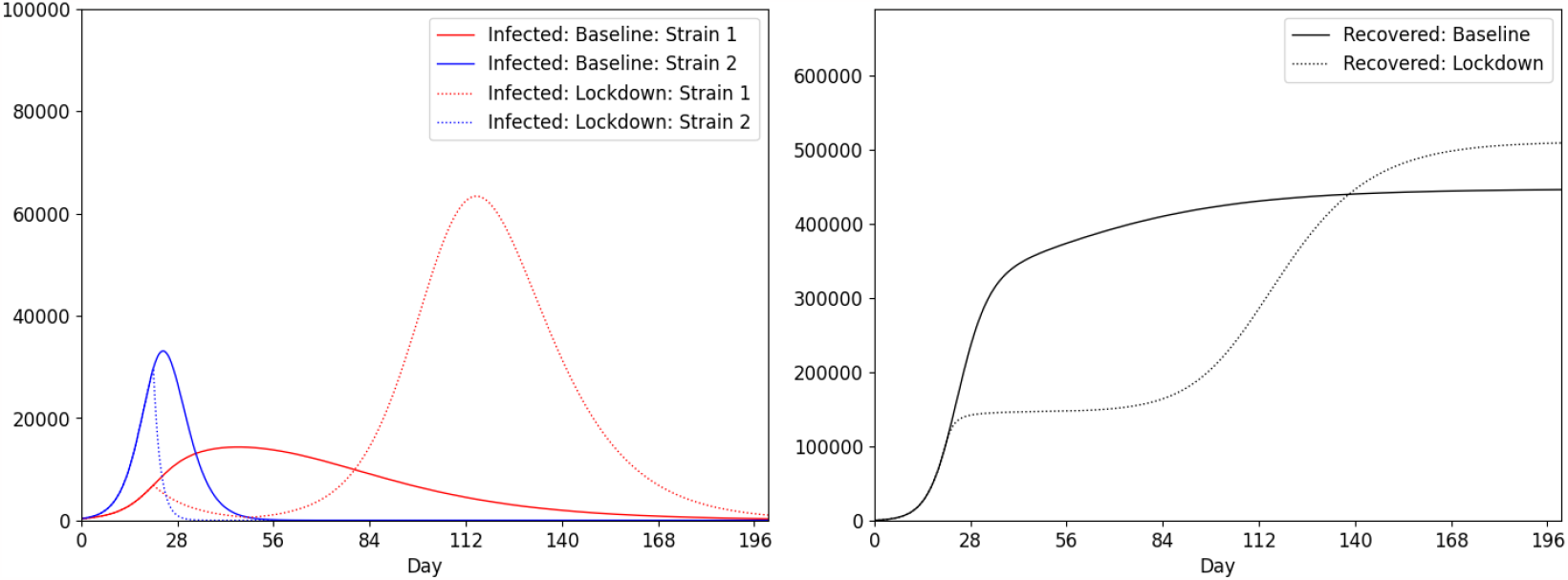
The impact of a lockdown in the two strain SIR model for (*a, b*) = (3.0, 0.2).

It is clear from Figure 2 that the lockdown, while temporarily reducing cases, later results a large second wave of **Strain 1** cases, thereby increasing the overall case count. Such effects are not visible in models that neglect the existence of multiple strains.

Moreover the existence of orange regions in Figure 1 is not sensitive to the initial ratio of cases between the two strains. Nor is it sensitive to the ratio of contact rates between the two scenarios, with the orange regions consistently appearing under variations of these parameters.

#### Lockdown Timing

Instead of fixing the start of the lockdown and varying the disease parameters, let us now fix the disease parameters and vary the start of the lockdown. Assuming one initial case for each strain, with now *β*_1_ = 0.35 and 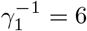 = 6 and ratios *a* = 3.6 and *b* = 0.2, we illustrate in Figure 3 the reduction in total cumulative cases resulting from a 28-day lockdown, with the lockdown starting on a range of different days. A positive reduction means the lockdown decreased total cases, while a negative reduction means the lockdown increased total cases.

**Figure 3:**
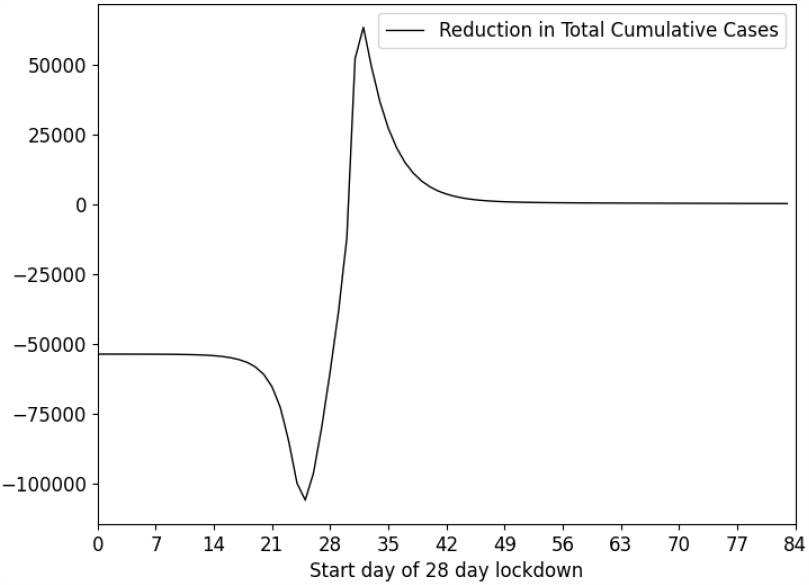
The impact of a 28-day lockdown in the two strain SIR model, with the lockdown starting on a range of different days.

Figure 3 shows that, for these particular parameters, the impact of the lockdown is very sensitive to timing. In particular, it shows that while starting the lockdown on day 33 results in the maximum reduction in cases, starting the lockdown *earlier* on day 25 results in a substantial increase in cases and the worst possible outcome. The dynamics of the system for these two start days are illustrated in Figure 4.

**Figure 4:**
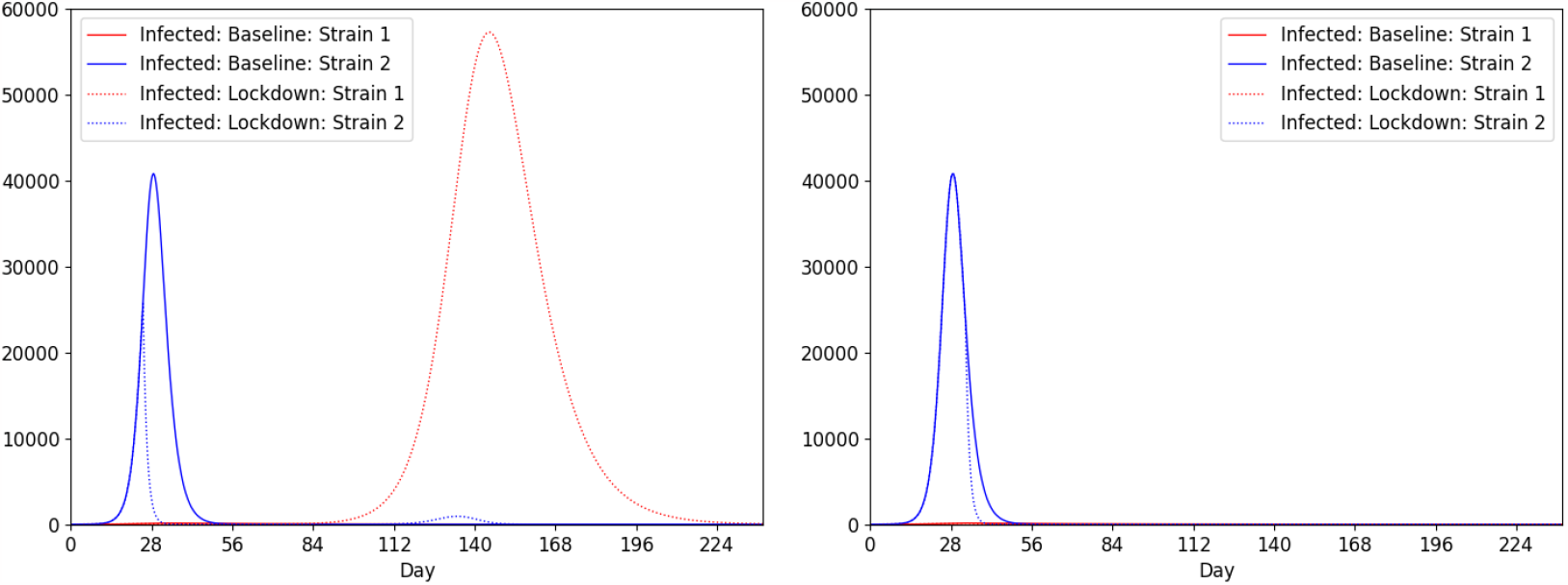
The impact of a 28-day lockdown in the two strain SIR model. On the left, the lockdown starts on day 25. On the right, the lockdown starts on day 33.

Figure 4 can be understood as follows. Starting the lockdown on day 25, **Strain 2** is quickly suppressed, giving **Strain 1** the advantage once the lockdown is lifted. On the other hand, starting the lockdown later on day 33 means that once the lockdown is lifted, sufficiently many individuals have already been infected with **Strain 2** that the population now has herd immunity against **Strain 1**, making the second wave impossible.

#### Multiple Regions

The situation is even more complex if multiple regions are involved. For example, given two identical copies of the population considered above, with a small amount of mixing between the two populations and with all initial cases located in only one region, the sensitivity to lockdown timing is even more pronounced, as illustrated in Figure 5.

**Figure 5:**
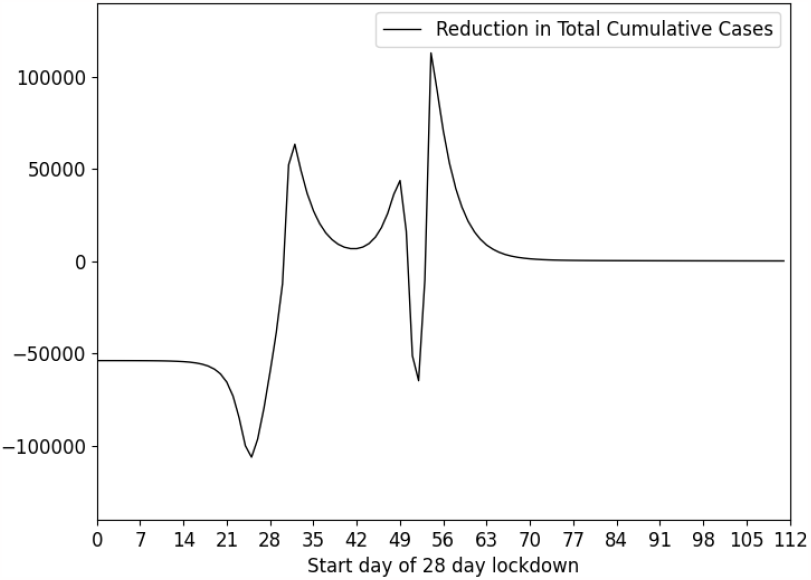
The impact of a 28-day lockdown in the two strain SIR model with two weakly interacting populations, with the lockdown starting on a range of different days.

A lockdown in one region might suppress the least transmissible variants there, allowing only the most transmissible ones to spread to neighbouring regions once the lockdown is lifted. A system consisting of multiple regions and multiple strains, with lockdowns for each region and travel restrictions for each pair of regions, with the timing of these interventions potentially variable, can be configured to produce even more undesirable outcomes.

### Agent-Based Model

We now turn to the agent-based model, where the lockdown can be implemented more realistically, to verify that our findings are not unique to the SIR model. In particular, we consider the following two scenarios:

- **Baseline:** No interventions are active.
- **Lockdown:** For a period of 28 days starting on day 21, individuals must stay at home unless in need of hospitalization.

We again consider the case of two strains, **Strain 1** and **Strain 2**, where as for the SIR model we assume that individuals cannot be simultaneously infected with both strains and that recovery from one strain implies immunity to both.

We randomly select equal numbers of initial cases for each strain. The reference **Strain 1** is configured using clinical monitoring data collected in Luxembourg between February 2020 and July 2020. The alternative **Strain 2** is configured by modifying the parameters of **Strain 1**. The transmission probabilities corresponding to **Strain 2** are given by multiplying the transmission probabilities of **Strain 1** by the *transmission probability ratio a*. The duration of time individuals spend in health states when infectious with **Strain 2** is given by multiplying the duration of time they would have spent in that health state, were they to have been infected with **Strain 1**, by the *infectious period ratio b*. We allow the pair (*a, b*) to vary over the same square (2) used for the two-strain SIR model, using a slightly coarser discretization, and for each pair (*a, b*) we plot the difference in total cumulative cases, after 400 days, between the **Baseline** and **Lockdown** scenarios, fixing the random seed for all simulations. The result is presented in Figure 6, using the same range and colour scheme as in Figure 1.

**Figure 6:**
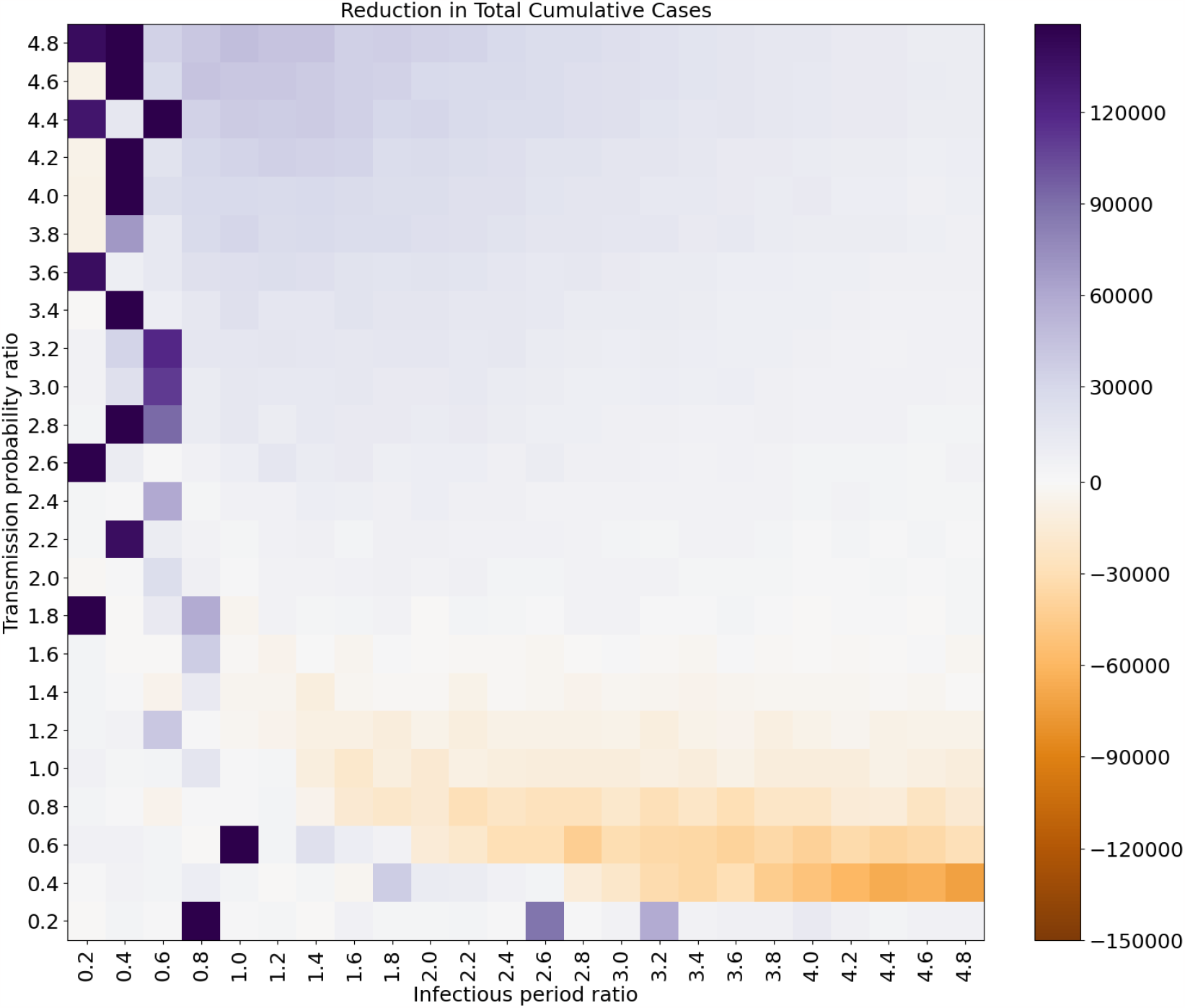
The impact of a lockdown in the two strain agent-based model. Orange regions are subsets of the parameter space in which the lockdown results in more cases than would have occurred without a lockdown.

Figure 6 is visually similar to Figure 1. In particular, using a agent-based model unrelated to the SIR model, we have verified the existence of orange coloured regions of the parameter space, where the lockdown increases total cases. In fact, the orange region in Figure 6 is broader than the equivalent region in Figure 1, indicating a larger subset of the parameter space containing unfavourable outcomes. Note that the dark purple squares in Figure 6, which cannot be found in Figure 1, correspond to simulations in which the virus was completely eradicated by the lockdown, something that is impossible in the SIR model.

In Figure 7 we plot daily cases and cumulative cases for the pair (*a, b*) = (0.6, 2.8). In this case, **Strain 1** has a higher transmission probability than **Strain 2**, while **Strain 2** has longer infectious periods.

**Figure 7:**
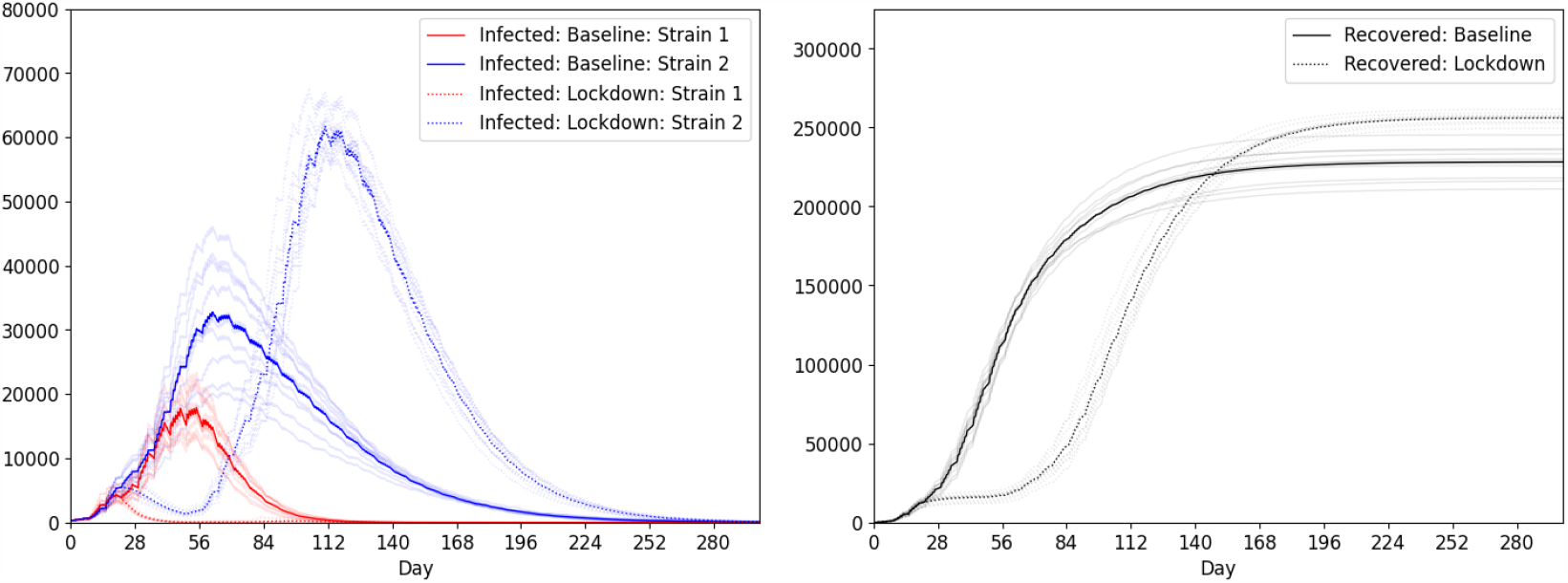
The impact of a 28-day lockdown in the two strain agent-based model for (*a, b*) = (0.6, 2.8).

As we saw earlier in Figure 2, we see in Figure 7 that the strain with the longer infectious periods is better able to endure the lockdown, and therefore gains a substantial advantage over the other strain once the lockdown is lifted, resulting in a large second wave and an overall increase in cases versus the baseline scenario.

## Discussion

Mutations to the SARS-CoV-2 virus have given rise to a large number of variants, with some of these variants believed to have the potential for increased transmissibility, increased virulence, or reduced effectiveness of vaccines. The biological fitness of these variants depends on their environment, including the behaviour of their host population. Throughout the COVID-19 pandemic, a range of non-pharmaceutical interventions have been deployed against the virus, including lockdowns, face masks, testing and contact tracing. These interventions have altered human behaviour and therefore had some impact on the competition between variants. In this article, we modelled the impact of a lockdown specifically and found that, under certain conditions, the competition between strains can be disrupted in such a way that ultimately leads to *more* infections than would have otherwise occurred without the lockdown. It is not, however, clear if under these conditions one would expect more hospitalizations and deaths, with this matter requiring further investigation.

Our results suggest that it would in practice be extremely difficult to implement a lockdown effectively, since necessary information on existing variants and their properties would typically be unavailable or incomplete. Without this information, it would be impossible to determine whether we are in the purple region in Fig 1, where the lockdown reduces cases, or in one of the orange regions, where the lockdown increases cases.

The results presented in this article are only the results of a modelling study, and we do not claim to have found evidence of these selection effects occurring in nature. That being said, during the first phase of the COVID-19 pandemic, the global response focussed on social distancing and other non-pharmaceutical interventions. It later shifted away from social distancing as vaccines became widely available. We might therefore speculate that the selective pressures exerted on the virus by the global response changed over time, and that these pressures may have played some role in the emergence of the Alpha and Delta variants and their eventual replacement by Omicron.

Putting together our results with those of Gurevich et al. [7], Ashby and Thompson [2] and Nielsen et al. [15], we conclude that interventions against COVID-19 might, under certain conditions, intensify the competition between variants in such a way that leads to a more resilient virus. Our conclusions may also apply to other viruses affected by the COVID-19 interventions, and more generally to any competition that may exist between them. For example, influenza strains now circulating in the human population may be more resilient to future non-pharmaceutical interventions, of the type implemented against COVID-19, than they would have been without the earlier interventions.

Our study is purely theoretical, and subject to numerous limitations. Our SIR model does not take into account hospitalization and death at all, while our agent-based model does not provide a realistic model of complex health care systems, neglecting for example their limited bed capacity. This is why we restricted our attention only to differences in total cases between the lockdown and baseline scenarios, as opposed to hospitalizations and deaths. Moreover our baseline scenario is somewhat unrealistic, since even in the absence of a lockdown individuals will, to certain extent, exercise self-protection and adjust their behaviour accordingly, leading to a narrowing of the difference between the baseline and lockdown scenarios.

The authors of the 2021 literature review by Jordan et al. [9], on optimization in the context of COVID-19 prediction and control, concluded that “a better understanding of the virus and its transmissibility, though challenging due to its evolutionary nature, would allow for more accurate and effective mitigation and resource allocation optimization efforts.” They also remarked that “better accessibility, uniformity and accuracy of data would provide vastly more robust and reliable mechanistic models, particularly when dealing with virus variants that continue to mutate and dampen mitigation efforts.”

A deeper and broader understanding of the impact of non-pharmaceutical interventions on the evolution of SARS-CoV-2 and other viruses could lead to greatly improved mitigation strategies in response to future pandemics.

## Data Availability

All data produced are available online at:
https://github.com/abm-covid-lux/multi_strain_abmlux_fast

## Acknowledgements

The authors would like thank Prof. Katrina Lythgoe of the University of Oxford for a very helpful discussion, and in particular for the interpretation of our results in terms of r/K selection theory.

## Notes

### Competing Interest Statement

The authors have declared no competing interest.

### Funding Statement

This study did not receive any funding.

